# A new Omicron lineage with Spike Y451H mutation that dominated a new COVID-19 wave in Kilifi, Coastal Kenya: March-May 2023

**DOI:** 10.1101/2023.07.03.23292158

**Authors:** Mike J Mwanga, Arnold W Lambisia, John Mwita Morobe, Nickson Murunga, Edidah Moraa, Leonard Ndwiga, Robinson Cheruiyot, Martin Mutunga, Laura M Guzman-Rincon, Charles Sande, Joseph Mwangangi, Philip Bejon, Lynette Isabella Ochola-Oyier, D James Nokes, Charles N Agoti, Joyce Nyiro, George Githinji

**Author notes:** Authors contributed equally to this article (Mike J Mwanga, Arnold W Lambisia, John Mwita Morobe, Nickson Murunga).

## Abstract

We report a newly emerged SARS-CoV-2 Omicron lineage, named FY.4, that has two unique mutations; spike:Y451H and ORF3a:P42L. FY.4 emergence has coincided with increased SARS-CoV-2 cases in coastal Kenya, April-May 2023. We demonstrate the value of continued SARS-CoV-2 genomic surveillance in the post-acute pandemic era in understanding new COVID-19 outbreaks.

## Main Text

To date over 340,000 test-confirmed COVID-19 cases and 5,688 COVID-19-related deaths have been reported in Kenya[1]. Sero-surveillance reports indicated a high seropositivity in rural and urban populations despite low vaccine uptake (27.9% of the adult population vaccinated with at least one dose)[2]. Specifically, by August 2022, 69%-81% of rural (Kilifi and Siaya) and 89%-95% of urban (Nairobi and Kisumu) adult population in Kenya had anti-Spike glycoprotein IgG antibodies (not published).

Genomic surveillance has been critical in informing origins of new waves, evolution, and geographical spread patterns of SARS-CoV-2. By June 2023, seven distinct waves of SARS-COV-2 infections were observed in Kenya[1][3]. The last three were dominated by Omicron sub-variants: BA.1-like, BA.5-like and BQ-like, respectively. These sub-variants were associated with increase in SARS-CoV-2 cases due to possession of mutations that conferred pre-existing immunity escape and transmission advantage[4].

In coastal Kenya, the KEMRI-Wellcome Trust Research Programme (KWTRP) has been conducting SARS-CoV-2 genomic surveillance across five health facilities (HF) within the Kilifi Health and Demographic Surveillance System (KHDSS)[5]. Samples are collected weekly from individuals of all ages presenting with acute respiratory illness (ARI) for screening. SARS-CoV-2 testing and sequencing is performed on: (i) samples collected from persons seeking medical attention from selected HF within the KHDSS and (ii) positive SARS-CoV-2 samples from collaborating public and private HF across Kenya.

Beginning late March, SARS-CoV-2 positivity rate in the HF increased from a background level of 1.2% in the week commencing 27^th^ March, and peaked at 42.9%, in the week commencing 24^th^ April (Figure, panel A). However, the positivity rate dropped in the first week of May to 23.5% and ranged between 5.0%-7.7% over the next three weeks. Between January and May 2023, 76 samples were sequenced and assigned into three lineages; BQ.1.1(n=1), BA.1.1(n=2) and FY.4(n=73). The increase in the positivity rate starting late March coincided with detection of a new Omicron lineage namely, FY.4 (Figure, panel B). In Kenya, the FY.4 lineage was first observed in Lamu County(n=4) on the 10^th^ March 2023 (Figure panel A). By 27^th^ May 2023, this new FY.4 lineage was further detected in four additional counties including the coastal counties of Kwale(n=2), and Mombasa(n=2), and central counties of Kiambu(n=5) and Nairobi(n=4). In Kilifi, FY.4 variants were first detected from samples collected on 27^th^ March 2023 and by April and May they became the predominant lineage representing 97% of all the detected lineages. Kenya is the first country to report the circulation of FY.4 lineage and has so far (based on GISAID data accessed on 30^th^ June 2023) been detected in eleven other countries; Austria, Germany, Italy, Sweden, Canada, France, China, Australia, Spain, United Kingdom and United States[6].

**Figure.**
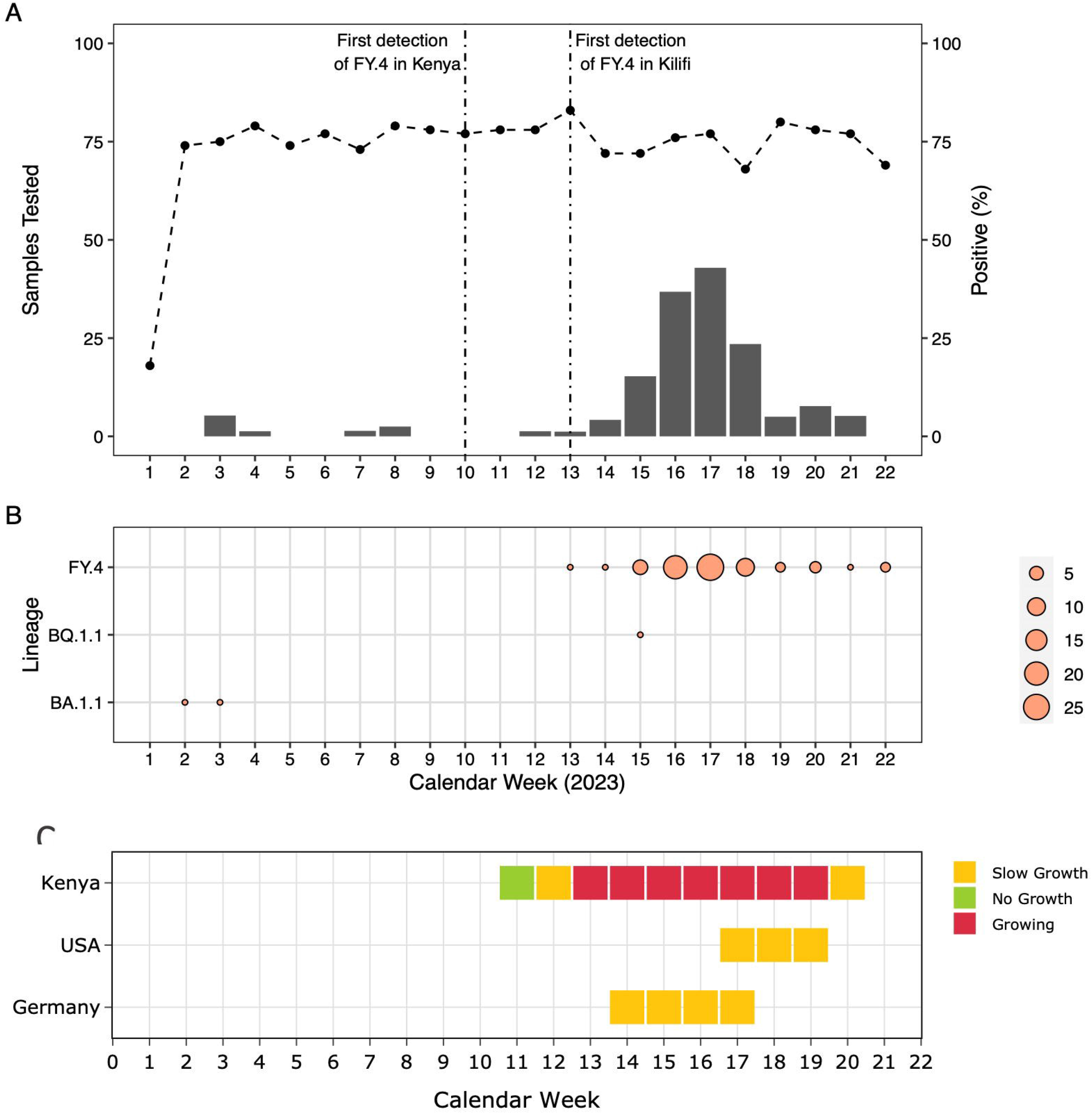
Panel A – Weekly number of collected samples (horizontal dotted line) and positive SARS-CoV-2 cases (bars) in health facilities within the Kilifi Health Demographic Surveillance System (KHDSS) between January – May 2023. Vertical dotted lines represent time points when FY.4 lineage was first detected in Kenya and in Kilifi. Panel B – Weekly distribution of SARS-CoV-2 lineages observed in the KHDSS between January – May 2023. Panel C – Growth rate estimates of the FY.4 variant in Kenya relative to USA and Germany.

Participants who had the FY.4 variant presented to the KHDSS HF mainly with cough (98%), fever (78%) and nasal discharge (74%) while 7% presented with difficulties in breathing (Table). Only 13 (16%) participants had received at least one dose of the AstraZeneca vaccine. However, a sero-surveillance study (February – June 2022) show that 67% of the unvaccinated KHDSS residents have anti-SARS-CoV-2 IgG antibodies showing that a high proportion of this population may have naturally acquired immunity from previous exposure[7].

**Table.** Distribution of observed clinical symptoms among the FY.4 cases observed in Kilifi Health Demographic Surveillance System between January – May 2023

| Symptoms | Omicron FY.4 (n=73) |
| --- | --- |
| Fever |  |
| Yes | 57 (78.1%) |
| No | 16 (21.9%) |
| Cough |  |
| Yes | 72 (98.6%) |
| No | 1 (1.4%) |
| Nasal discharge |  |
| Yes | 54 (74.0%) |
| No | 19 (26.0%) |
| Difficulty in breathing |  |
| Yes | 5 (6.8%) |
| No | 68 (93.2%) |
| Sore throat |  |
| Yes | 28 (38.4%) |
| No | 45 (61.6%) |
| Body malaise |  |
| Yes | 25 (34.2%) |
| No | 48 (65.8%) |
| Conscious level |  |
| Alert | 73 (100.0%) |
| COVID-19 Vaccination status |  |
| Yes | 12 (16.4%) |
| No | 60 (82.2%) |
| No data | 1 (1.4%) |
| COVID19 vaccine doses |  |
| 1 | 3 (4.1%) |
| 2 | 7 (9.6%) |
| No data | 63 (86.3%) |

Relative to other Omicron lineages, the FY.4 has two additional amino acid mutations; in the spike(Y451H) and ORF3a(P42L) genes. The potential phenotypic impact of the Y451H change remains unknown. Previous studies have shown that spike amino acid change in the receptor binding domain (RBD) near the Y451H, such as L452R increases virus infectivity and fusogenicity by enhancing spike stability and cleavage[8]. However, mutations within the ORF3a CD8^+^ T cell epitopes have been reported to cause complete loss of recognition in the ancestral lineages and Alpha VOC[9].

We applied a Bayesian hierarchical model[10] to estimate the growth rate of the FY.4-like lineage in Kenya. These estimates serve as warning system for lineages showing consistent increase in frequency for at least two consecutive weeks in Kenya and/or other countries. Growth rate estimates on Kenyan data was compared to data from Germany and USA, as these were the only countries with reported FY.4 cases in at least two consecutive weeks as of the last weeks of May (Figure, panel C). The model has warned of a high level of concern in Kenya as from week of March 26 towards May, suggesting continued increase in cases attributed to the FY.4 lineage.

In summary, surveillance of SARS-CoV-2 in Kenya has detected the emergence of a new Omicron lineage with unique spike and ORF3a gene mutations. Detection of FY.4 lineage coincided with increase in SARS-CoV-2 cases in Kilifi and has also been detected in other parts of the country. Growth estimates suggests potential for continued increase in geographical spread of FY.4. Further analysis on the immunological impacts of the observed mutations and any transmission advantage arising are ongoing.

## Supporting information

gisaid_supplemental_table_epi_set_230627zw.pdf

## Data Availability

Genome sequences generates in this study are available on GISAID. Generated genomes are listed in the gisaid_supplemental_table_epi_set_230627zw.pdf. The, the dataset and analysis scripts used are available in Havard Dataverse at https://doi.org/10.7910/DVN/ZMGR5P.

## Acknowledgement

We thank the laboratories, hospitals and organisations that shared specimens for sequencing at KWTRP and the submitting laboratories where genetic sequence data were generated and shared via the GISAID Initiative, on which this research is based. Submitting and the Originating laboratories of the GISAID data used in this study are listed in the gisaid_supplemental_table_epi_set_230627qs.pdf

## Ethical Statement

The whole genome sequencing study protocol was reviewed and approved by the Scientific and Ethics Review Committee (SERU) residing at the Kenya Medical Research Institute (KEMRI) headquarters in Nairobi (SERU # 4035).

